# Temporal DCE Profile of Brain Metastasis with A Comparison of Pseudoprogression Cases

**DOI:** 10.1101/2022.12.19.22283618

**Authors:** Sevcan Turk, Ryo Kurokawa, Shotaro Naganawa, Jacob Wallace, Tianwen Ma, Timothy Johnson, Toshio Moritani, John Kim

**Author notes:** Corresponding Author; 1500 E. Medical Center Drive, UHB2-A209, Ann Arbor, MI 48109-5030. Presented at RSNA 2021. An IRB-exempt approval was obtained for this retrospective study from the University of Michigan.

## Abstract

**Purpose:** To demonstrate Dynamic Contrast-Enhanced (DCE) perfusion changes in brain metastasis patients after chemoradiation therapy within the treatment responders (true-response group and pseudoprogression group) and between the true-response and pseudoprogression groups.

**Materials and Methods:** 38 true-response patients with 38 brain metastases (13 melanoma, 11 lung, 7 breast, 7 others) with 3 consecutive DCE-MRI examinations (pretreatment, first follow-up and second follow-up) and 7 pseudoprogression patients with 10 melanoma metastases with 2 consecutive DCE-MRI examinations (pretreatment, first follow-up) were evaluated. DCE-MRI parameters and permeability graphs increase (rapid, slow, medium) and course pattern (washout, plateau, persistent) were analyzed and compared between the timing of the examinations and between the true-response and pseudoprogression groups, using paired t-tests and Mann-Whitney U-tests as appropriate. Automatic assessment of washout curves by Olea Brain Software was reviewed.

**Results:** Pretreatment mean wash-in (15.3±24.7 vs 6.50±14.2, P<0.02), Vp (0.13±0.17 vs 0.06±0.06, P<0.01), Ktrans (0.37±0.48 vs 0.16±0.18, P<0.01), peak enhancement (123±98 vs 76±60.5, P<0.01), AUC (14×10^4^±18×10^4^ vs 5×10^4^±10×10^4^, P<0.001) values were significantly higher than that of post-treatment. Other DCE metrics and permeability graph factors between pre-and post-treatment MRI were not significantly different. There was no statistically significant difference in any imaging factors in pre-or first post-treatment DCE-MRI examinations between the true-response and pseudoprogression groups.

**Conclusion:** Wash-in, Vp, Ktrans, peak enhancement, AUC values in DCE perfusion were significantly decreased after chemoradiation therapy in the responders. No statistically significant difference was shown between the true-response and pseudoprogression groups on either pre-or post-treatment, indicating the homogeneous permeability profiles in the responders.

## Background/Purpose

Secondary brain neoplasms or brain metastases are defined as intra-axial lesions metastasizing from tumors outside the central nervous system (CNS) and are the most common cause of malignant brain tumors in adults. Chemotherapy and radiotherapy of brain metastases are the mainstay of treatment. Treatment response assessment is crucial for the management, but may be complicated by pseudoprogression, an increase in contrast-enhancing lesion size followed by improvement or stabilization without any further treatment, from the first few weeks to 6 months after completing treatment (1). Significant effort has been devoted to distinguish responders (true-response and pseudoprogression) and non-responders (true-progression) in brain metastases using various imaging biomarkers. After treatment four different imaging patterns may be seen; rapid decrease in size, stable disease with late treatment response, initial increase with delayed tumor regression, or new lesions followed by a decrease in size of the overall tumor burden (7). The last two patterns reflect pseudoprogression. However, if the responder is a homogeneous group or not has not been proven, although these previous studies have assumed it is without proof.

Dynamic contrast-enhanced (DCE) MRI is a perfusion MRI method that has been used to assess lesions in the brain, head and neck, and spine. DCE perfusion provides information about contrast agent leak through abnormal tumor capillaries and other contrast dynamics within the tumor tissue with contrast-time curves (4). Quantitative DCE parameters are Vp (plasma volume), Ve (extracellular space volume), Ktrans (transfer volume constant from plasma to tissue), and Kep (transfer rate constant). Semiquantitave parameters are derived from permeability curves which are AUC (area under the curve), peak-enhancement (maximum contrast concentration), wash-in (enhancement rate), wash-out (de-enhancement rate), SER (signal-enhancement ratio), TME (time to maximum enhancement) (5). The main advantages of extended Tofts-Kety Model DCE parameter calculations are Ktrans, Vp, and Ve show accurate permeability and/or plasma flow values that are weakly vascularized or highly perfused (4).

In general, malignant tumors show a high and fast enhancement followed by wash-out, whilst benign tissues show a more progressive enhancement (4). A recent study compared the quantitative DCE parameters before and after radiotherapy in brain metastases between non-responders and responders, and found that pre-treatment Ve and Ktrans were significantly higher in non-responders (8). We considered that it is essential to evaluate the presence or absence of the difference in DCE parameters between the true-response and pseudoprogression groups.

Our aim is to investigate the chronological changes of quantitative DCE MRI parameters in brain metastases after chemoradiation therapy and compare the parameters between the true-response and pseudoprogression groups.

## Materials and Methods

### Study Population

An institutional review board approval was obtained for this retrospective study. Informed consent was waived. A single institutional database search with the Electronic Medical Record Search Engine (EMERSE) was done to generate a list of intracranial metastasis patients who received chemoradiation therapy and presented to our Radiology department.

165 patients with brain metastasis who received chemoradiotherapy were reviewed from late 2017 to early 2021. 120 patients were excluded due to lack of pre-treatment or two consecutive follow-up MRIs with DCE perfusion images or due to small lesions (<3 mm) which were not measurable on perfusion images. Treatment response and pseudoprogression were decided based on consecutive at least 3 follow up MRIs. Pertinent medical records including clinical notes, surgical and pathology reports were reviewed and correlated with imaging findings.

Finally, forty-five patients with brain metastasis were extracted from the list. Among those, thirty-eight patients with brain metastasis (13 melanoma, 11 lung, 7 breast, 7 others) were classified as true-response group. They underwent pretreatment, first follow-up (approximately 3^rd^ month after treatment) and second follow-up (approximately 6^th^ month after treatment) brain MRIs including DCE perfusion examinations. The other seven patients with 10 metastatic melanomas were classified as pseudoprogression group. They underwent pretreatment and first follow-up MRIs.

### Data Acquisition

All conventional brain MRIs, DWI and DCE MRI sequences were acquired with 5 mm slice thickness and 1 mm gap using 1.5T or 3T MRI machines (Achieva 1.5T, Ingenia 1.5T and Ingenia 3T; Philips Health Systems, Eindhoven, The Netherlands). Axial T1W, axial T2W FLAIR, sagittal pre and postcontrast 3D T1 TFE, sagittal 3D BrainView axial DWI and additionally DCE MRI were performed using 3D T1-weighted fast-field echo images, with the administration of 20 mL gadobenate dimeglumine contrast agent (MultiHance; Bracco Diagnostics) with 4.0 mL/s flow rate.

The parameters for pre and postcontrast 3D T1W TFE sequence were as follows: FOV: 240 mm, slice thickness/gap: 1/-mm, voxel: 1×1×1 mm, TR:shortest. Postcontrast sagittal T1W 3D BrainView TSE sequence parameters were FOV: 250 mm, slice thickness/gap: 1.1/-mm, voxel: 1×1×1 mm, TR:600. The DWI EPI sequence parameters were FOV: 240 mm, slice thickness/gap: 5/1 mm, voxel: 1.5×1.8, TR: shortest. DCE perfusion was a FFE sequence with FOV: 200 mm, slice thickness/gap: 5/-mm, voxel: 1×1×1 mm, TR: shortest. Eight and 16 channel head coils were used for the acquisition.

### Data Analysis

Region of interests (ROIs) were manually placed covering the solid enhancing region of the tumor without necrosis or hemorrhage on post-contrast axial T1-weighted image by three neuroradiology fellows under the observation of two expert neuroradiologists with more than 10 and 30 years of experience in image interpretation, respectively. Mean perfusion parameters within the ROIs were automatically calculated using Olea Sphere 3.0 (Olea Medical Solutions, La Ciotat, France) Brain Suite Permeability Module. Washin-washout graphs with increase type and course of the signal intensity time curve were automatically created by the software using time-intensity curves according to Tofts-Kety model.

All quantitative DCE MRI parameters include SER, wash-in, wash-out, Kep, Vp, Ve, Ktrans, TME, peak, curve-washout, peak-enhancement, and AUC. Automatic assessment of washin-washout graphs including increase type (rapid, slow, or medium) and course pattern (washout, plateau, or persistent) of the graphs were obtained by using Olea Brain Suite and given as percentages (Figures 1 and 2).

**Figure 1:**
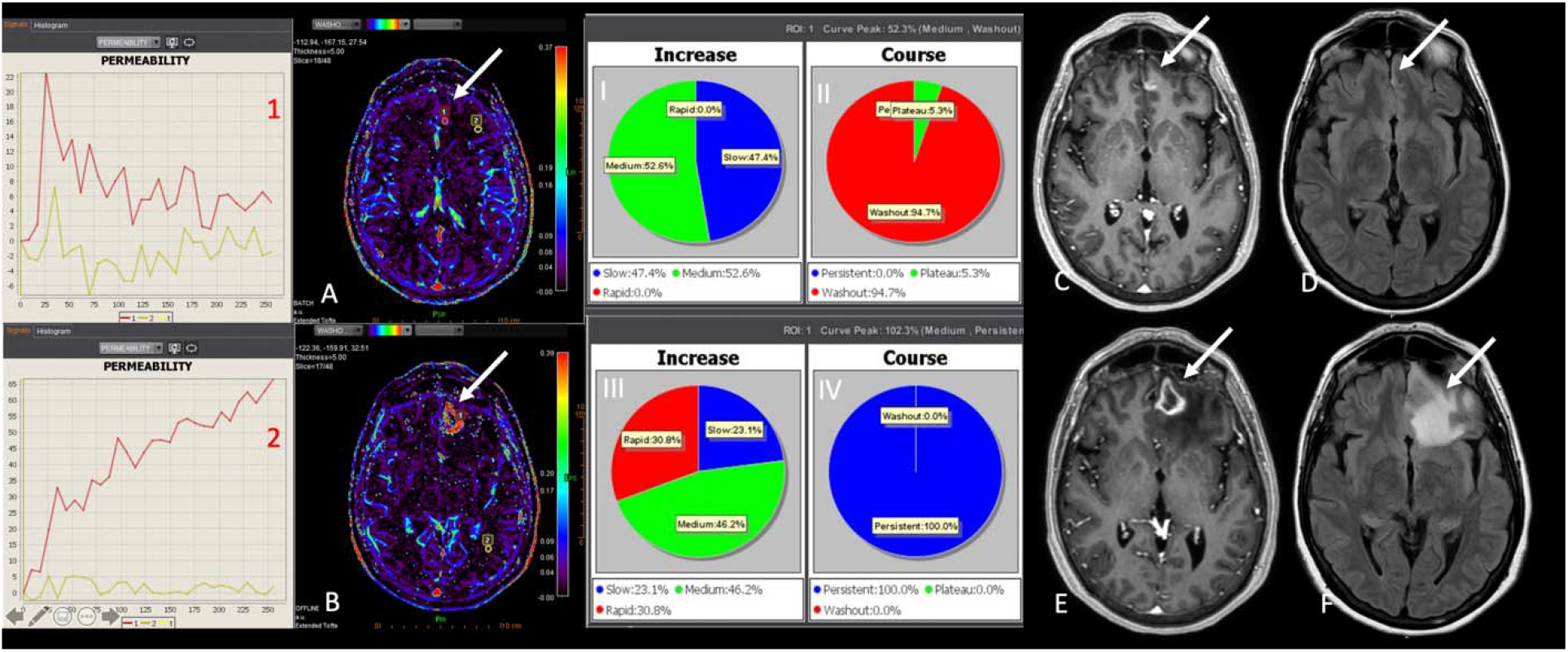
Lung cancer metastasis to left anterior frontal lobe. Upper row images are belong to pretreatment study. The DCE Washout maps with relatively medium increase pattern with washout course of the permeability graph. Lower low images show changes after chemoradiation. Enhancing lesion size and perilesional edema increased. Washout DCE map shows elevated signal, however permeability graph course show persistent pattern with medium increase, suggestive for pseudopragression. (Follow up images showed decreased in size which is consistent with pseudoprogression.)

**Figure 2:**
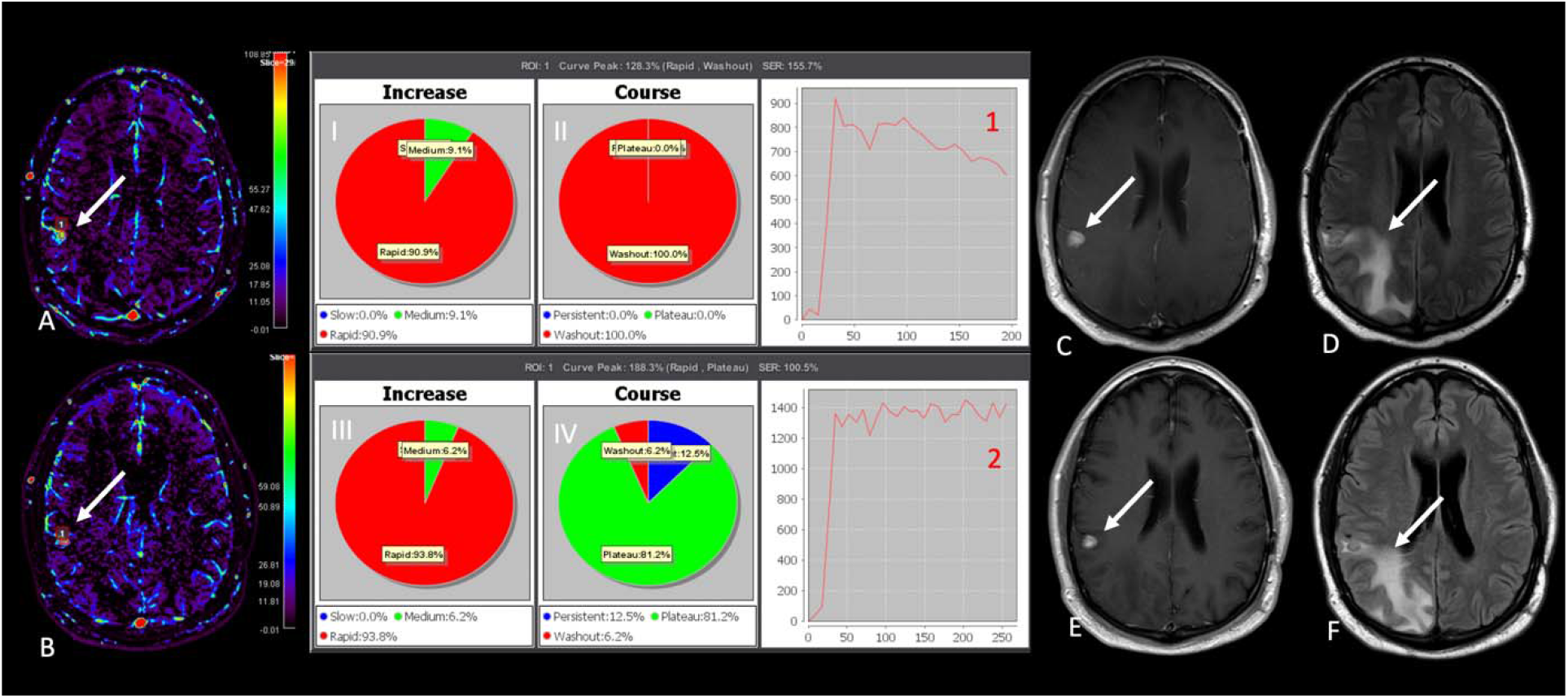
Right posterior frontal melanoma metastasis, Upper row shows pretreatment images with washout course of the perfusion wash-in map with rapid increase. Lower row shows early images after chemoradiation. Enhancing lesion slightly decreased in size with slightly increased perilesional edema. Wash-in perfusion map shows visually no significant change, however, plateau course pattern with rapid increase which are suggestive for treatment response. (Follow up images showed prominent decrease in size consistent with treatment response)

### Statistical Analysis

Mean values of DCE parameters (SER, wash-in, wash-out, KEP, VP, VE, Ktrans, TME, peak, curve-washout, peak-enhancement, and AUC) were compared between pre-treatment and the first follow-up MRIs, between the first and second follow-up MRIs, and between the true-response and pseudoprogression groups using paired t-test, Kruskall-Wallis, Mann Whitney U test. The Shapiro-Wilk tests were performed to test the normality of the continuous variables. To compare permeability graph course and increase Pearson’s chi-squared tests were performed to test the heterogeneity given the contingency tables. Raw p-values were adjusted by false discovery rate due to multiple comparisons.

Analysis was performed with SPSS and Python SciPy software. P < 0.05 was considered statistically significant.

## Results

Mean age was 60.5 ± 12 (females mean age: 62 and male mean age 60) years old. Mean time between pre-treatment and the first follow-up MRI studies was 95 days, and between the first and second follow-up MRI studies was 97 days.

Pretreatment mean wash-in (15.3 ± 24.7 vs 6.50 ± 14.2, P<0.02), Vp (0.13 ± 0.17 vs 0.06 ± 0.06, P<0.01), Ktrans (0.37 ± 0.48 vs 0.16 ± 0.18, P<0.01), peak enhancement (123 ± 98 vs 76 ± 60.5, P<0.01), AUC (14×10^4^ ± 18×10^4^ vs 5×10^4^ ±10×10^4^, P<0.001) values were significantly higher than that of post-treatment (Table 1).

**Table 1:** Comparison of pre and post-treatment DCE parameters.

| **DCE**<br>**PERFUSION**<br>**PARAMETER** | **PRE-**<br>**TREATMENT**<br>**MEAN VALUE**<br>**(± SD)** | **POST-**<br>**TREATMENT**<br>**MEAN**<br>**VALUE (± SD)** | **P-**<br>**VALUE**<br>**(adjusted)** |
| --- | --- | --- | --- |
| SER | 126 (116) | 130 (135) | 0.49 |
| **WASH-IN** | **15 (24)** | **6.5 (14.2)** | **0.016** |
| WASH-OUT | 6.5 (13.6) | 8.8 (18.9) | 0.73 |
| KEP | 1.4 (2.3) | 0.89 (0.84) | 0.36 |
| **VP** | **0.1 (0.17)** | **0.06 (0.06)** | **0.01** |
| VE | -0.13 (2.69) | 0.26 (0.25) | 0.33 |
| **K-TRANS** | **0.37 (0.48)** | **0.16 (0.18)** | **0.01** |
| TME | 181 (238) | 160 (78) | 0.37 |
| **PEAK** | **712 (852)** | **299 (391)** | **0.06** |
| CURVE<br>WASHOUT | 7.1 (26.3) | 14.9 (14.0) | 0.23 |
| **PEAK**<br>**ENHANCEMENT** | **123 (98)** | **76.0 (60.5)** | **0.01** |
| **AUC** | **14x10<sup>4</sup> (18x10<sup>4</sup>)** | **5x10<sup>4</sup> (7x10<sup>4</sup>)** | **0.00** |

There was no statistically significant difference between pre- and post-treatment SER, washout, Kep, Ve, TME, peak, or curve washout values. There was no statistically significant difference between the first and second follow-up DCE parameters for true-response groups. There was no statistically significant difference between the true-response and pseudoprogression groups in any DCE perfusion parameters, permeability graph increase, or course.

Pre-treatment lesions tended to have more washout type graphs with rapid increase whereas post-treatment lesions tended to show persistent or plateau graph course with slow increase pattern on the signal intensity time curve graphs (Figures 3 and 4).

**Figure 3:**
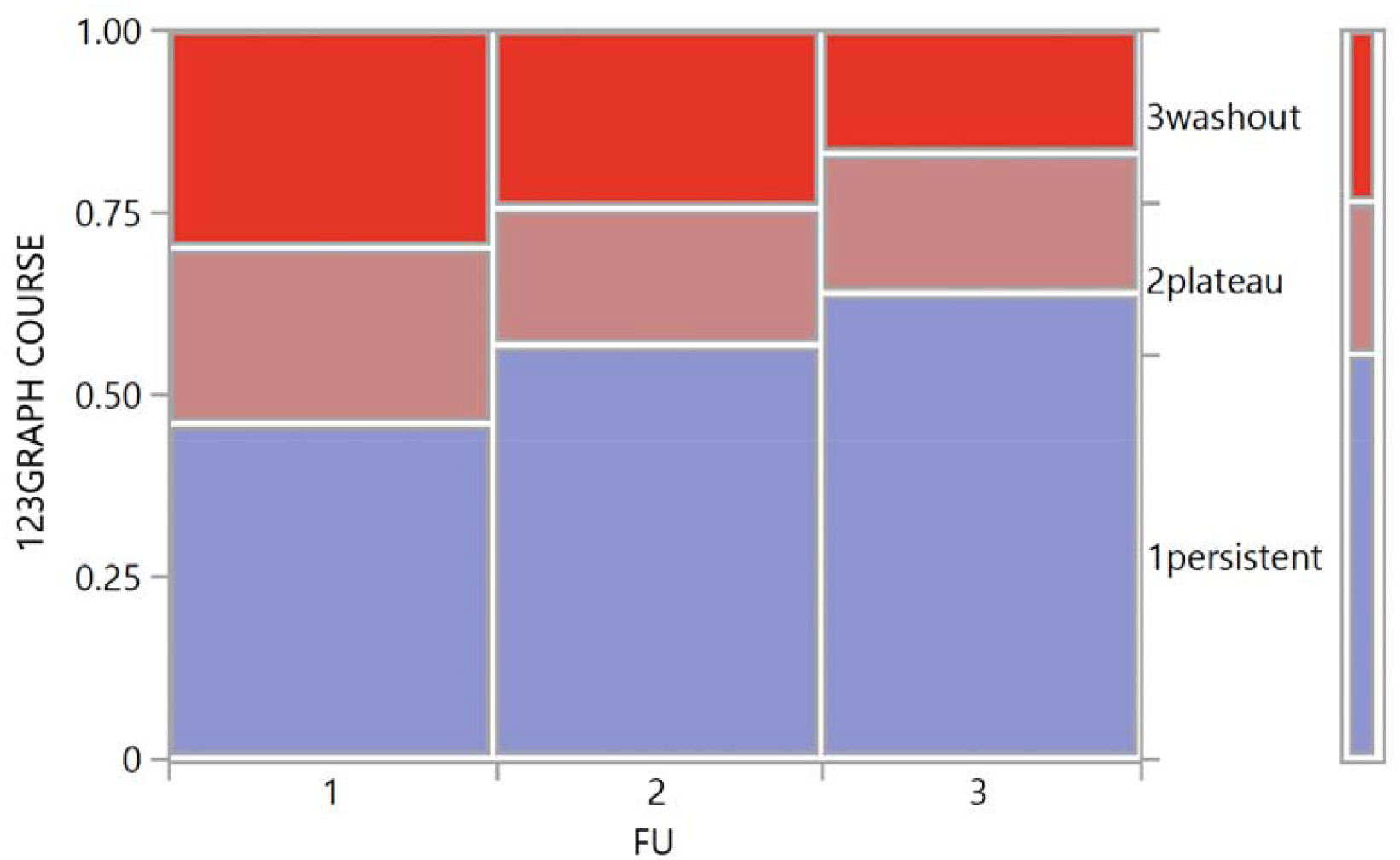
Graph course of permeability maps in all 3 follow-up MRIs in the true-reseponse group; pre-treatment, the first follow-up, and the second follow-up.

**Figure 4:**
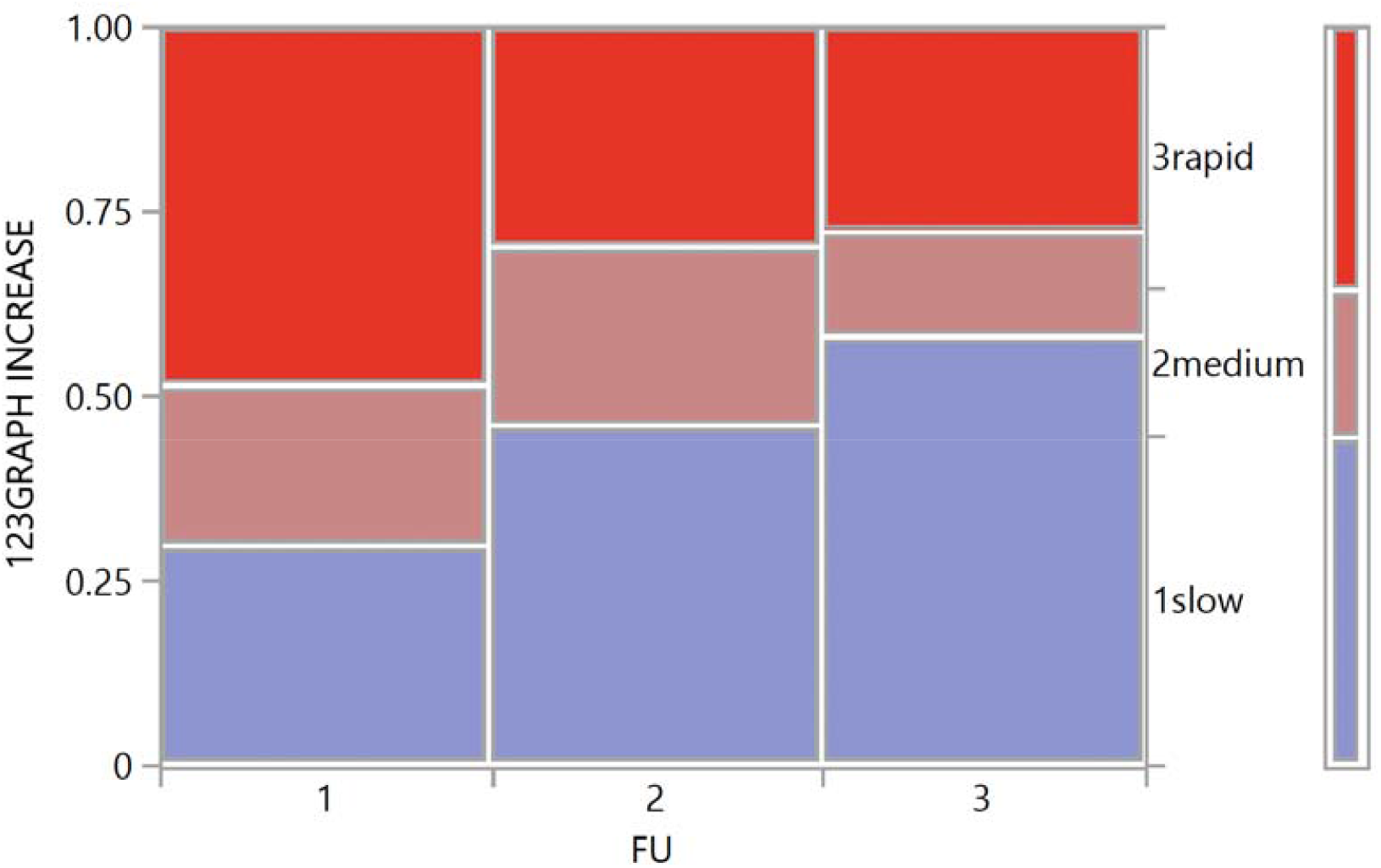
Graph increase of permeability maps in all 3 follow-up MRIs in the true-response group; pre-treatment, the first follow-up, and the second follow-up.

## Discussion

This study evaluated the pre-and post-treatment DCE perfusion parameters within and between the responders (true-response and pseudoprogression groups) of brain metastasis treated with chemoradiotherapy. We found statistically significant differences in multiple DCE-MRI parameters between pre-and post-treatment examinations. Post-treatment lesions tended to have a slow increase and plateau or persistent type graph, compared with the pre-treatment lesions. We also found no significant difference in the DCE-MRI parameters between the true-response and pseudoprogression groups, suggestive of the homogeneity of the permeability profiles within the responders.

DCE-MRI could be a promising tool to show tumors’ leakiness and vascular microenvironment. Parameters in DCE-MRI have been reported to be useful in differentiation between the true-and pseudoprogression in patients with high-grade gliomas (9,10). However, the literature is very limited for DCE parameters in brain metastasis and treatment-associated changes in those patients. Morabito et al showed that Ktrans was significantly decreased in radiation necrosis compared with the true-progression group in 72 intra-axial tumors (11). However, they combined the primary brain tumors and metastases, and they did not measure any other DCE parameters or their chronological changes. Hatzoglou et al. reported significantly decreased Vp and Ktrans in radiation injury compared with true-progression group in 26 patients with metastases (14), but they did not evaluate the chronological changes, either. Furthermore, the patterns of the permeability graphs have not been established in brain metastasis regardless of the response to treatment.

In this study, post-treatment true-response and pseudoprogression lesions tended to show persistent or plateau patterns with more slow increase when compared to pre-treatment DCE permeability graphs. Kuhl et al showed that rapid washout graph pattern was associated with malignant lesions while benign neoplasms demonstrated slow persistent and plateau course (12). Jung et al showed that permeability graph AUC was useful to differentiate melanoma metastasis and glioblastoma from hypovascular brain metastasis (13). They also concluded that glioblastoma and hypervascular melanoma metastasis tended to have rapid washout graph course whereas hypovascular brain metastasis tended to have plateau and persistent courses (13). Rapid increase and washout pattern has been considered to be associated with an increased tumor capillary vessels and their enhanced leakiness (13). The changes in the trends of the DCE permeability graphs after treatment in the present study indicated the reduced tumor capillary vascularity and leakiness, possibly represented by the decreased wash-in, Vp, and Ktrans on the postrreatment MRIs.

Studies of treatment-associated changes in DCE parameters in brain metastasis have been scarce. Moreover, the difference in DCE parameters between the true-response and pseudoprogression groups has been poorly understood, although these two groups can be regarded as the single “responder” group in comparison to the non-responder group [ref]. On conventional MRI sequences, the tumor becomes small in size and less enhanced in the true-response group, whereas pseudoprogression refers to MRI findings mimicking true tumor progression without significant tumor cells. Pseudoprogression lesions primarily consist of treatment-induced necrosis, inflammatory cells, post-operative infarcts, post-ictal changes, and increased vascular permeability, which cause increase in size of enhancement (15). Discrimination of pseudoprogression is challenging and depends on decrease in size on the follow up MRIs. Shah et al analyzed DCE parameters among brain metastasis before and 72 hours after radiotherapy in 16 patients with 25 lesions, and demonstrated significantly higher Ve and Ktrans in non-responders compared with responders (8). However, to our best knowledge, DCE parameter comparison between the true-response and pseudoprogression groups has not been focused on. There was no significant DCE parameter difference between the two groups, indicating the homogeneous nature of pseudoprogression and post-treatment groups.

Limitations of the study include a small number of subjects to compare the lesions with heterogenous primary tumors. This is only single institution’s experience. ROI was considered to represent the whole tumor, however, tumor heterogeneity, hemorrhage, necrosis may alter the perfusion parameters. Small lesions (<3mm) were not visible on perfusion maps which were excluded from the analysis.

More research can be done to separate between treatment responders and pseudoprogression from true progressive disease. Signal intensity time curve graphs may provide additional information beyond the evaluation of high or low perfusion parameters. Some peudoprogression cases may show elevated perfusion signal however the course type of the signal intensity time curve especially for wash-in maps may be helpful to discriminate pseudoprogression from true tumor progression.

## Conclusion

We have revealed the natural course of DCE perfusion parameters after chemoradiation treatment in responders, including the true-response and pseudoprogression groups. No significant difference among the responders indicated the homogeneity of the responders and potentially reinforce the robustness of the research method of comparing the DCE parameters between responders and non-responders.

Contributions and Acknowledgements

K.M.L. and J.C.C. conceived the study. K.M.L. and C.A.T. performed nucleic acid extraction and library preparation for population samples. K.E.G., J.A., C.R.P., and J.C.G. provided field and colony collections. K.M.L. performed genome assembly and population analyses. K.M.L. and J.C.C. wrote the manuscript with input from all authors.

Thank you to Robert Munch (QB3, UC Berkeley) and John Alterio (PacBio, Menlo Park) for their advice and work on DNA extraction, library preparation, and long-read genome sequencing for the genome assembly. Thank you to Dr. Graham Coop and Dr. Jeffrey Ross-Ibarra for providing advice on population analyses.

This work is funded by NIFA 2020-67013-30976 and NIFA CA-D-ENM-2150-H.

## Data Availability

All data produced in the present study are available upon reasonable request to the authors

## Abbreviations

DCE Perfusion: Dynamic contrast enhanced perfusion
Vp: Plasma volume
Ve: Extracellular space volume
Ktrans: Transfer volume constant from plasma to tissue
Kep: Transfer rate constant
SER: Signal enhancement ratio
AUC: Area under the curve

## Notes

### Competing Interest Statement

The authors have declared no competing interest.

### Funding Statement

no external funding was received

### Author Declarations

An IRB-exempt approval was obtained for this retrospective study under the HUM00193148 number from the University of Michigan.

